# The Feasibility of Smartwatch Micro Ecological Momentary Assessment for Tracking Eating Patterns of Malaysian Children and Adolescents in the SEACO Child Health Update 2020: a Cross-Sectional Study

**DOI:** 10.1101/2025.08.12.25333193

**Authors:** Richard Lane, Louise A. C. Millard, Ruth Salway, Sophia M. Brady, Andy L Skinner, Chris J. Stone, Jeevitha Mariapun, Sutha Rajakumar, Amutha Ramadas, Hussein Rizal, Laura Johnson, Tin Tin Su, Miranda Elaine Glynis Armstrong

## Abstract

**Background:** Mobile-phone ecological momentary assessment (EMA) methods are a well-established measure of eating and drinking behaviours, but compliance can be poor. Micro-EMA (μEMA), which collects information with single tap response to brief questions on smartwatches, offers a novel application that may improve response rates. To our knowledge, there is no data evaluating μEMA to measure eating habits in children or in low-to-middle income countries.

**Objective:** We investigated the feasibility of micro-EMA to measure eating patterns in Malaysian children and adolescents.

**Methods:** We invited 100 children and adolescents aged 7-18 in Segamat, Malaysia to participate in 2021-2022. Smartwatches were distributed to 83 children and adolescents who agreed to participate. Participants were asked to wear the smartwatch for 8 days and respond to 12 prompts hourly from 8am to 8pm, asking for information on their meals, snacks and drinks consumed. A questionnaire captured their experiences using the smartwatch and μEMA interface. Response rate (proportion of prompts responded to) assessed participants’ adherence. We explored associations between response rate with time of day, across days, age and sex using multi-level binomial logistic regression modelling.

**Results:** Eighty-two participants provided usable smartwatch data. The median number (inter-quartile range) of meals, drinks and snacks per day were 2 (2 - 4), 3 (1 - 5) and 1 (0 - 2) respectively on the first day of the study. The median response rate across the study was 68% (quartiles: [50, 83]). The response rate decreased across study days from 74% (68, 78) on day 1 to 40% (30, 50) on day 7 (odds ratio [OR] per study day: 0.73 [95% confidence interval [CI]: 0.64, 0.83]). Response rate was lowest at the start of the day, and highest between the hours of 12:00-14:00. Female participants responded to more prompts than male participants (OR: 1.72 95% CI: [1.03, 2.86). There was no evidence of differential response by age (OR: 0.73 95% CI: [0.41, 1.28]). Most participants (65%) rated their experience using the smartwatch positively, with 33% saying they were happy to participate in future studies using the smartwatch. For children that didn’t wear the smartwatch for the full study duration (n=22), discomfort was the most common complaint (41%).

**Conclusions:** In this study of the feasibility of μEMA on smartwatches to measure eating in Malaysian children we found the method was acceptable. However, response rates declined across study days resulting in substantial missingness. Future studies (e.g. through focus groups) should explore approaches to improving response to event prompts, trial alternative devices to increase children’s comfort and evaluate revised protocols for reporting of intake events.

## Introduction

Non-communicable diseases (NCDs) are the most common cause of death worldwide [1]. In Malaysia [2] NCDs particularly impact lower-income households [3]. Therefore, health surveillance in this population is required to better understand policy interventions that may improve health outcomes in Malaysia. Dietary risk factors accounted for 10% of all deaths globally in 2021 [4], therefore, measuring eating is a crucial component of health surveillance. Traditional methods for measuring eating and dietary intake include food diaries, 24 hour recalls, diet histories or food frequency questionnaires. While methods relying on memory of past behaviour are subject to error like recall bias, prospective methods like diaries are affected by reactivity, where real or reported behaviour is altered owing to the process of documenting food intake in real-time. Under-reporting is common in all existing methods, with an estimated 263 kcal/day typically missing from self-reported intakes compared with objective measures [5]. Under-reporting is differential by type of food and eating event with snacks and snack foods more likely to be left out of a self-reported record compared with direct observation [6] [7], or data from wearable cameras [8] [9]. Online tools, such as Intake24, that guides users through a 24-hour recall process aims to reduce researcher burden in coding data collected. Photographic methods, where participants are asked to take pictures of their meals rather than write down each food and drink along with its portion size, aim to offer a more objective approach to add portion size estimation and reduced participant burden for capturing real-time food intake [10] [11] [12]. However, moving 24 hour recall online has not yet altered estimated under-reporting [13] [14], and issues with remembering to take photos before consuming foods as well as automating the estimation of foods and nutrients [15] in photos mean that outstanding challenges in dietary assessment methods remain [16].

Ecological Momentary Assessment (EMA) is the repeated sampling of current behaviours in real-time in a natural environment [17]. EMA has evolved to be primarily delivered using mobile phones (mEMA), which have improved response rates compared with original pen and paper methods [18] [19] [20]. There is a large volume of literature on EMA using smartphones (n=796 studies) [21]. While diet is the second most commonly studied topic it still only accounts for 4% (n=35) of these studies. Studies of diet using EMA in young people are primarily in US and Europe, with just two studies in Asia, in China and Taiwan [19] [22].

Liao [23] highlighted that responses rates and compliance with EMA protocols were rarely reported. Since then, reporting of compliance has improved, but the response latency remains unknown from many studies [22]. Response rates to mEMA of diet are median 74% [24], which is similar to mEMA of all topics (mean 75% (IQR 64%, 84%) [21]. A review of mEMA for diet in young people (16-30) showed response rates mostly exceeded 80% [22], whereas at younger ages poorer responses <80% are more often observed [19]. Lower response rates have also been associated with weekend vs. weekdays [22]; when participants receive more prompts during the day [18] [21] and in males vs. females [22].

Smartwatches are an emerging technology for collecting data alongside sensor data using micro EMA (μEMA) protocols. This captures information using single tap responses to brief questions which is suitable for the small screens on these devices [24] [20]. In children and adolescents, the use of pen and paper or mobile EMA to measure diet has typically been implemented outside of school hours [19]. Smartwatches offer the potential to implement EMA across the whole day. To our knowledge, only two diet studies involving adults in the UK and US have reported on EMA with smartwatches, and none have involved children [24] [25] [19]. μEMA has been found to yield higher compliance [26]. Further, the use of μEMA significantly improves response rate (mean 72% vs. 82%) but remains rare with only 12 studies on any topic in any age group [21].

Therefore, this study investigates the feasibility of using smartwatch-based μEMA to record eating patterns in Malaysian children and adolescents. The collected EMA data is used to examine the completeness of the collected data, factors associated with response rates, alongside survey responses assessing participants’ experience during the study.

## Methods

### SEACO-CH20 Study

The South-East Asian Community Observatory (SEACO) Health and Demographic Surveillance cohort is a dynamic cohort of 13,335 households in Segamat, a semi-rural region in the state of Johor Darul Takzim, Malaysia. The cohort was established in 2012, with surveys, blood tests, and physical measurement data collected from participants. In 2013 and 2018, health surveys were conducted on ∼25,000 adults and children, 25,168 in 2013 and 24,710 in 2018.

Children and adolescents aged 7-18 years who were part of the SEACO cohort were invited to participate in the SEACO Child Health 2020 update (SEACO-CH20) study; a systematic review of EMA studies in youth recommended 7 as a lower age limit for EMA [27]. The eligibility of households was limited by location due to the safety measures implemented during the COVID-19 pandemic to reduce the risk to participants, households and fieldworkers. Therefore, the 1,993 children and adolescents invited to participate were from only 3 of the 5 SEACO sub-districts (Jabi, Sungai Segamat and Gemereh) in the Segamat district.

Data collection visits were performed in person from 1st November 2021 to 31st July 2022. The data collected included surveys, physical measurements such as height, weight, blood pressure, waist and hip circumference and blood sample collection. Participants were given wrist-worn Axivity AX6 6-axis accelerometers to monitor their activity. A subset of the participants were also given TicWatch C2 Android smartwatches to record eating and drinking with μEMA, using a smartwatch μEMA system developed within the research team. The smartwatch system used was an adaptation of a µEMA system first used in a study involving high-temporal density longitudinal measurement of alcohol use [28] by a subset of the research team who developed this system. Participants were briefed on the use of these devices by the data collectors.

Written informed consent was obtained from parents or guardians on behalf of the participants. Children and adolescents were also asked to provide their written assent to participate in the study. Ethical approval was obtained from the Monash University Human Research Ethics Committee on 17/03/2020 (Project ID: 23271) and University of Bristol REC Case no. 2020 – 4208(ID nr.: 1304255).

### Study Participants

Participants for the SEACO-CH20 study were selected from the larger SEACO cohort. Parents of participants provided consent for their children to participate in SEACO-CH20. A sub-sample of 100 participants were each invited to wear a smartwatch.

### Data Collection

SEACO-CH20 fieldworkers performed two home visits to collect the data. The smartwatch was distributed on the initial visit, and the participants were briefed on how to use and charge it. They were instructed to wear the device for the next 8 days, on “the wrist that [they] use to write”. The smartwatches were collected during the second home visit, and the participants were asked to complete a questionnaire on their experience with the devices. Questionnaires assessed the participants’ attitudes to several aspects of the smartwatch study including ease of use, their attitude toward charging and their overall experience. This was based on similar pilot work using novel methods in the ALSPAC G2 study [29] and included the following questions:

- Overall, how would you rate your experience of using the smartwatch during the study, on a scale from 1 (didn’t like it at all) to 5 (really liked it)?
- If you were asked to use the smartwatch again in another study, would you participate?
- How many days in total did you wear the smartwatch for?
- If you wore the smartwatch for less than 8 days, what were the main reasons for not wearing it longer?

Parents of participants aged 7-9 years completed the survey on behalf of their children, while participants aged 10 and older filled out the survey themselves

### Smartwatch μEMA questions

During the study, the smartwatch prompted participants once every hour to enter any food or drink that they had consumed in the last hour. These prompts were scheduled to appear once every hour from 9am to 8pm, so participants were expected to interact with the smartwatch 12 times throughout the day. The smartwatch interface included the following five questions that the participants completed for each item consumed:

- Have you had any food or drink in the last hour? Options: Yes; no
- What did you have? Options: Meal; snack; drink
- What size was it? Options: Small; medium; large
- What did you use to eat? Options: Hands; fork/spoon; chopsticks
- Where were you? Options: Home; school; elsewhere

After entering this information for one item consumed, they were asked “Any more food or drink to record?” and could then start again to add another entry. Therefore, each consumption entry either indicates that the participant did not eat or drink in the last hour, or contains the answers to the above questions for a particular meal, drink or snack, linked to an hour period within a day. If participants ignored the prompts, they would receive a reminder prompt after 1 minute; if they continued to ignore the prompt for a further 1 minute the prompt would disappear and “no response” would be recorded by the smartwatch.

Participant could choose ‘back’ on each question screen to return to a previous question and update their response. However, after submitting their answers for a particular item (i.e., completing the ‘where were you’ question for that item), they would not be able to return to that entry.

An additional prompt (the “catch-up”) was scheduled every morning at 8am asking if they had consumed any food or drink on the previous day that had not been recorded on the smartwatch. If they indicated ‘yes’, they were asked the same questions as above. Catch-up entries did not have an associated eating time but were labelled as catch-up type events, indicating that they applied to the previous day.

The smartwatch study was co-created and piloted with the Malaysian research team and the English translated into Malay for use on the smartwatch. All the original data collection was in Malay.

### Smartwatch Data Cleaning

Smartwatches were distributed by fieldworkers partway through the day, and μEMA responses on this distribution day were removed from analyses. The study period is taken to be the subsequent seven days after this distribution day.

The version of the EMA software we used did not save the hour period to which each entry belonged. Therefore, we needed to infer this from the submission timestamp – the date and time a particular entry was submitted. As entries for the same hour period are submitted one after the other, we used a time window to group nearby entries into a single “eating event”, which is intended to capture the participants’ responses to one prompt. A 30-minute window was chosen to group nearby prompts, as we expect this to collect entries from the same eating event without grouping prompts from adjacent hours. Previous work from diet diaries suggests that 30 minutes is a reasonable cut-point to distinguish independent eating occasions [30]. Occasionally, there may be participants with more than 12 eating events per day; if, for example, they took more than 30 minutes to finish responding to a prompt. This occurred on 26 occasions, less than 5% of the total 574 (82 participants ^*^ 7 days) study days.

The μEMA data used was restricted to the 7 days after the distribution day. During the data review, we identified an issue with the collected data where there were sometimes multiple identical entries for a given intake event due to an issue with the μEMA software. Therefore, duplicate entries were identified as any pair of entries with identical contents (same meal type, portion size, utensil, and location), for the same hour period, and entered within five minutes of each other. The first such entry was kept in each case. 588 duplicate responses were removed of 10,539.

Data cleaning was performed in Python version 3.10.0.

### Response Rate

The response rate was calculated as the proportion of prompts responded to (with either at least one item consumed or an entry stating that they did not eat or drink anything in the previous hour). The response rate tells us the extent that participants engaged with the smartwatch app throughout the day but not the extent that the data entered is complete, i.e., whether all intake events were recorded. We therefore summarised the number of each type of meal entry (meal, drink, snack, or no food/drink) submitted per day across our sample. For these summaries we included only participants who took part in the study outside the Ramadan fasting period (3rd April– 1st May 2022, n = 67), since fasting participants are likely to enter fewer eating events during the day. The mean of participants’ response rates per day was recorded, and the median and quartiles of these reported.

Attrition from the study was examined by identifying the last day each participant responded to any smartwatch prompt – participants who had ceased responding to the prompts are referred to as “inactive”.

#### Statistical Analyses

We summarised response rates to each individual prompt of the smart-watch μEMA and the participants’ experiences based on the survey questions. We used mean for continuous variables, n % for categorical variables and median and interquartile range (IQR) for ordinal or non-normally distributed continuous variables.

We used a mixed-effect logistic regression model for the response (yes/no) to an individual prompt on a specific study day of data collection for each participant. A fixed effect term was included for study day (from the first to the seventh day) as a continuous linear trend. The time of day was also included as a fixed effect to capture nonlinearity in response throughout the day, grouping the prompts by the nearest hour as follows to decrease the number of parameters in the model:

- Morning (09:00 – 11:00)
- Lunchtime (12:00 – 14:00)
- Afternoon (15:00 – 17:00)
- Evening (18:00 – 20:00)

Random intercept and random slope terms were included for study day within each participant. Estimates are provided as odds ratios (OR), and 95% confidence intervals (95% CI), interpreted as the multiplicative change in the odds of a participant responding to an individual prompt. The degree of difference between participants was summarised in the intraclass correlation coefficient.

To evaluate if changes in participation across wear days differed in boys versus girls, we repeated this base model adding a fixed term for sex and an interaction term between sex and study day. Similarly, we explored differences by age group (Malaysian primary school age: 7-12 years versus secondary school age: 13-18 years) by adding a fixed term for age group and an interaction term between age group and study day to the base model.

Analyses were performed in Python version 3.10.0 and R version 4.2.2 [31]. All of our analysis code is publicly available [32]. Git tag 3.0 corresponds to the version of the analyses presented here.

## Results

### Participants

A flow chart showing the study participants can be seen in Figure 1. Parents of 728 participants consented to their children’s participation in SEACO-CH20 of which 615 consented to wear the accelerometer. Of those who consented, 100 participants were invited to wear the smartwatch. Of the 100 participants invited to participate in the smartwatch sub-study, 83 participants agreed. **Error! Reference source not found**. The reasons for non-participation included concern the device was not comfortable (N=3) and allergies (N=2). The remaining participants rejected the smartwatch without comment. One further participant accepted the smartwatch study but removed the EMA app from the watch during the study period, rendering their data unrecoverable, resulting in 82 participants who provided smartwatch data.

**Figure 1.**
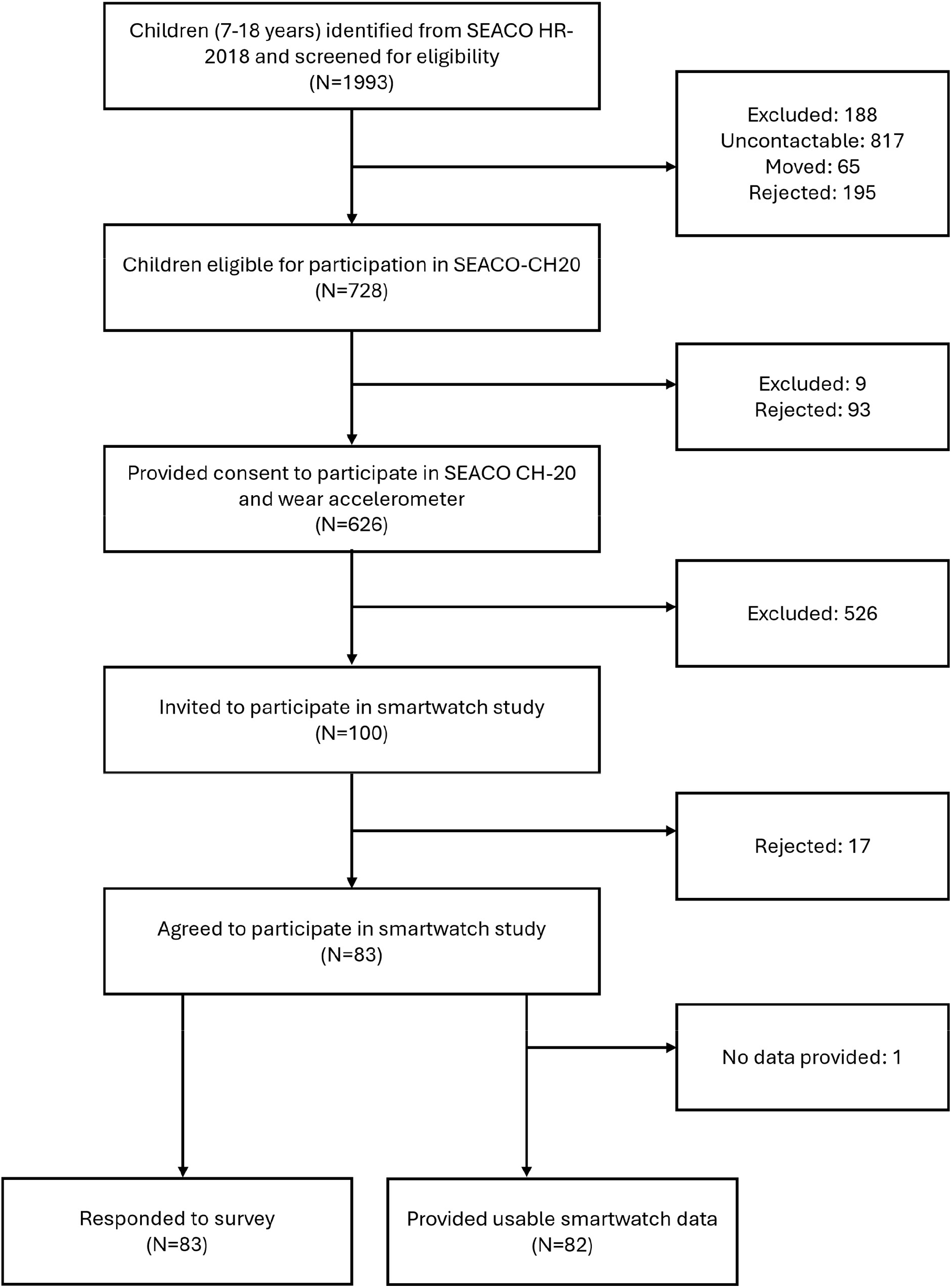
Study flow chart. Reasons for rejecting the smartwatch study included concern about discomfort and allergies.

The sex, ethnicity and age breakdown for all participants who took part in the smartwatch study can be seen in Table 1.

**Table 1.** Summary of participant demographics (N=83).

|  |  | Smartwatch participants |  |
| --- | --- | --- | --- |
| Participant characteristic | Category | N (83) | % |
| Sex | Female | 53 | 64% |
|  | Male | 30 | 36% |
| Ethnicity | Malay | 73 | 88% |
|  | Non-Malay | 10 | 12% |
| Age (years) | 7-12 | 24 | 29% |
|  | 13-15 | 28 | 34% |
|  | 16-18 | 31 | 37% |

### Smartwatch Responses

The median prompt response rate was 69% (IQR: [52, 82]%).

The number of participants who became inactive on each day can be seen in **Error! Reference source not found**.. The majority (55/82, 67%) of participants were active until day 7 – i.e., they responded to at least 1 prompt on day 7.

The median and IQR in the number of entries of each type made by each participant per day is summarised in Table 2. Fifteen participants took part (at least partially) during Ramadan and so were excluded from these summaries. Only a minority of intake events were submitted as catch-up entries (N=125 catch-up entries versus 4705 non-catch-up).

**Table 2.**
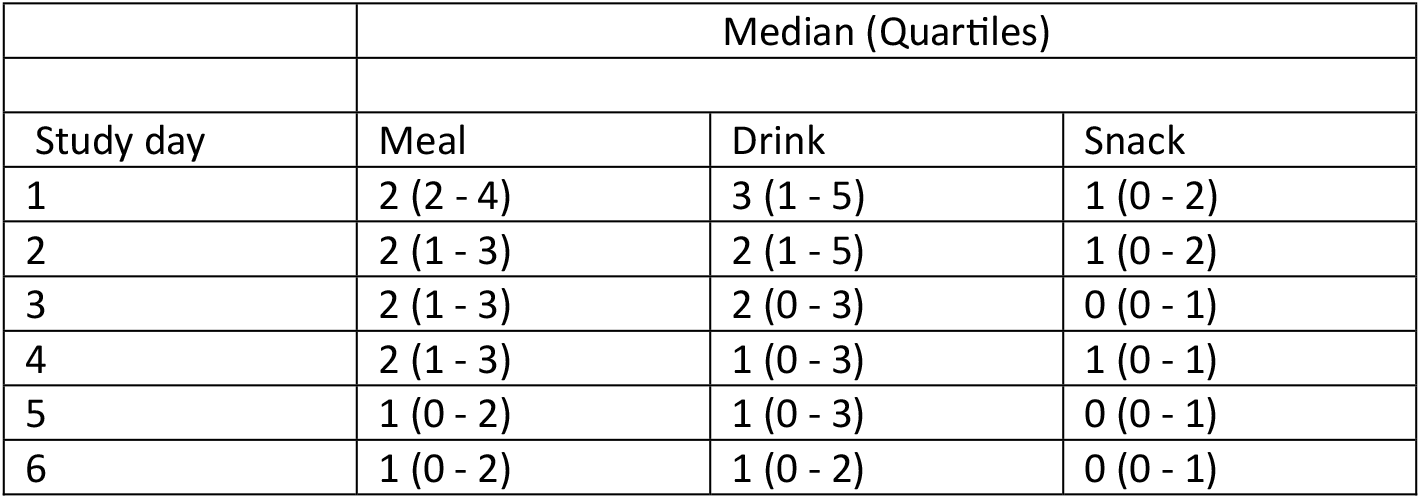
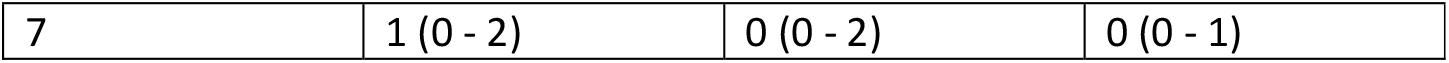
The median and interquartile range of the number of non-catch-up entries per day per participant, for participants whose study period did not intersect with Ramadan (n=67).

#### Response Rate across and within study days

The response rate for individual prompts had a median (IQR) of 67% (50, 83). The response rate on each day ranged from 83% (66-92) on day 1 to 58% (33-75) on day 7.

Figure 3 shows the response rate with study day and time. The response rate decreased across study days (OR for each additional day of the study: 0.73 (95% CI: 0.64, 0.83). The response rate was lowest at the beginning of the day; the OR and 95% CI are summarised in Table 3. The intraclass correlation coefficient was 0.207, which indicates that approximately 21% of the total variance in prompt response behaviour was attributable to between-participant differences.

**Table 3.** the odds ratios for responses at different times of day, taking breakfast time (9-11am) as the reference level. The response rate was lowest at the beginning of the day.

|  | Odds ratio | 95% CI |
| --- | --- | --- |
| Breakfast (09:00 – 11:00) | 1.00 (Reference level) | - |
| Lunchtime (12:00 – 14:00) | 1.69 | [1.43, 2.12] |
| Afternoon (15:00 – 17:00) | 1.49 | [1.25, 1.76] |
| Evening (18:00 – 20:00) | 1.27 | [1.07, 1.51] |

**Table 4.**
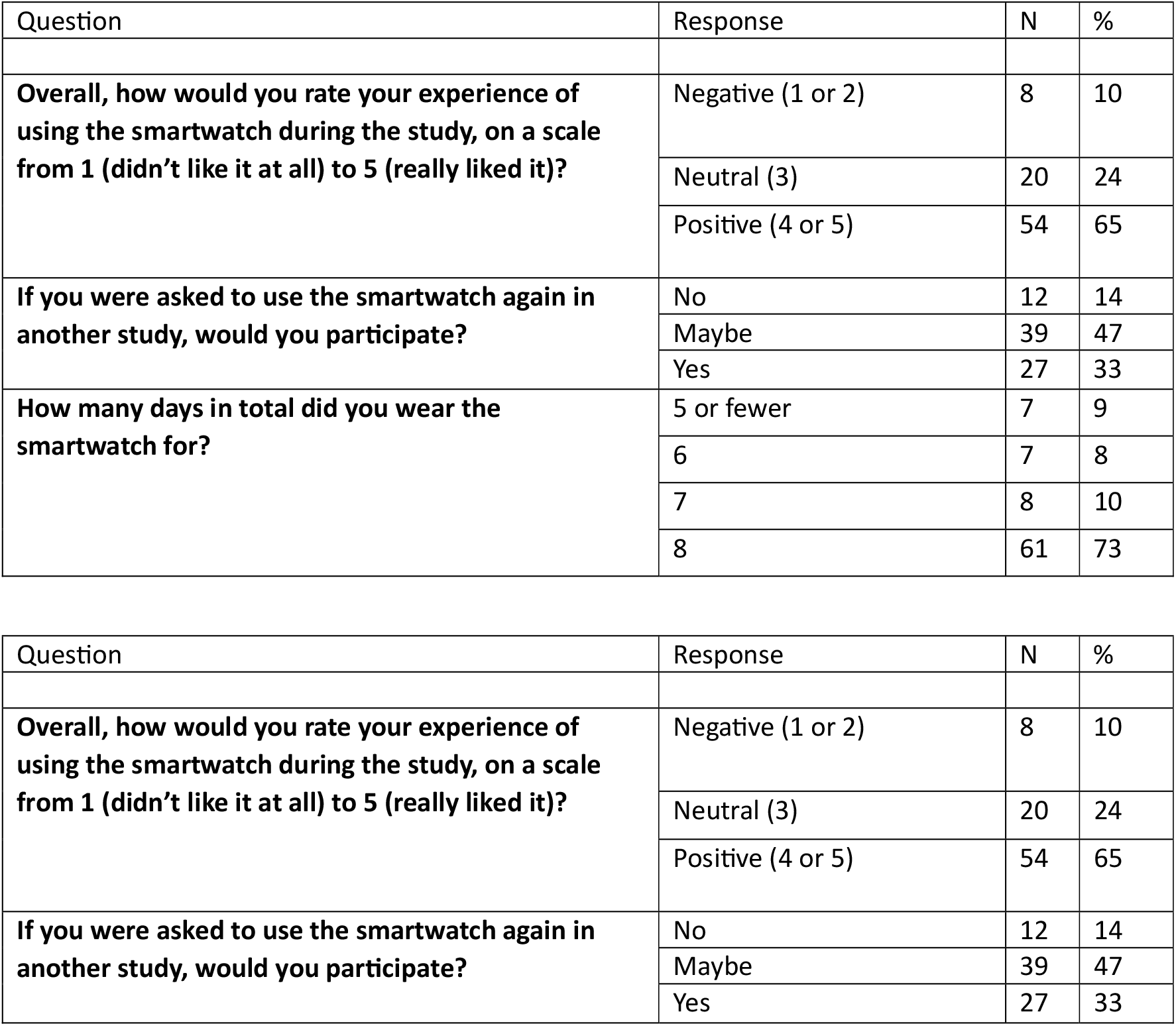
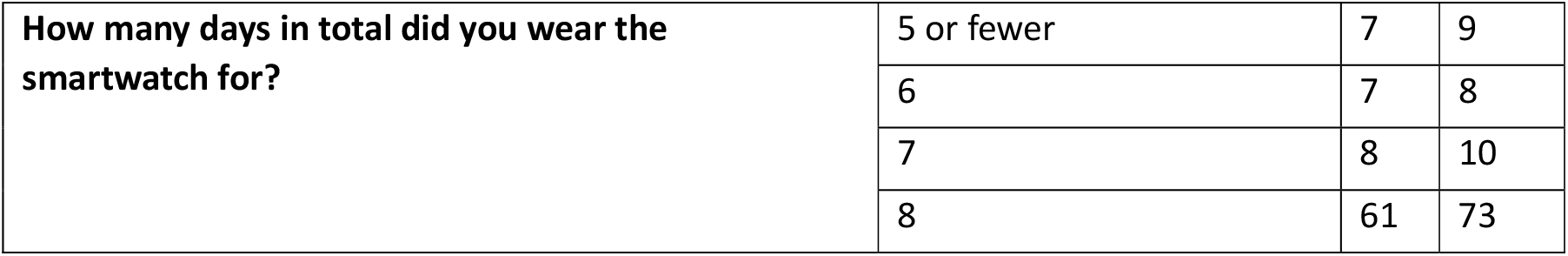
The survey questions. Participants were directed to wear the smartwatch on the day that the watch was distributed and for the seven subsequent days making 8 days total. Missing data and participants who refused to respond are not included. N=83.

| Question | Response | N | % |
| --- | --- | --- | --- |
| **Overall, how would you rate your experience of using the smartwatch during the study, on a scale from 1 (didn't like it at all) to 5 (really liked it)?** | Negative (1 or 2) | 8 | 10 |
|  | Neutral (3) | 20 | 24 |
|  | Positive (4 or 5) | 54 | 65 |
| **If you were asked to use the smartwatch again in another study, would you participate?** | No | 12 | 14 |
|  | Maybe | 39 | 47 |
|  | Yes | 27 | 33 |
| **How many days in total did you wear the smartwatch for?** | 5 or fewer | 7 | 9 |
|  | 6 | 7 | 8 |
|  | 7 | 8 | 10 |
|  | 8 | 61 | 73 |

| Question | Response | N | % |
| --- | --- | --- | --- |
| **Overall, how would you rate your experience of using the smartwatch during the study, on a scale from 1 (didn't like it at all) to 5 (really liked it)?** | Negative (1 or 2) | 8 | 10 |
|  | Neutral (3) | 20 | 24 |
|  | Positive (4 or 5) | 54 | 65 |
| **If you were asked to use the smartwatch again in another study, would you participate?** | No | 12 | 14 |
|  | Maybe | 39 | 47 |
|  | Yes | 27 | 33 |
| **How many days in total did you wear the smartwatch for?** | 5 or fewer | 7 | 9 |
|  | 6 | 7 | 8 |
|  | 7 | 8 | 10 |
|  | 8 | 61 | 73 |

**Figure 2.**
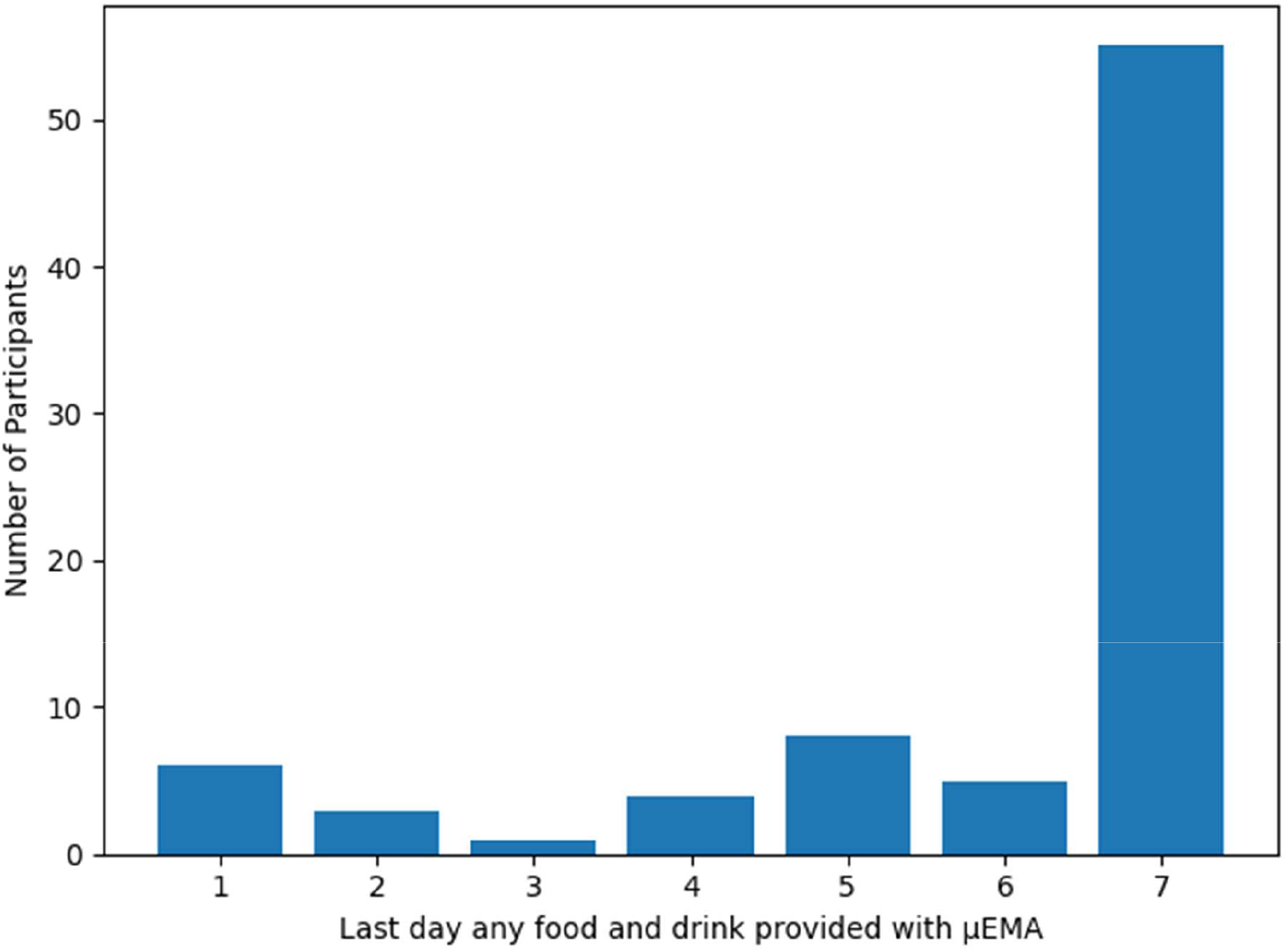
The number of participants who became inactive on each day (N=82), i.e., having responded to no μEMA prompts after this day. All participants were active for at least one day.

**Figure 3.**
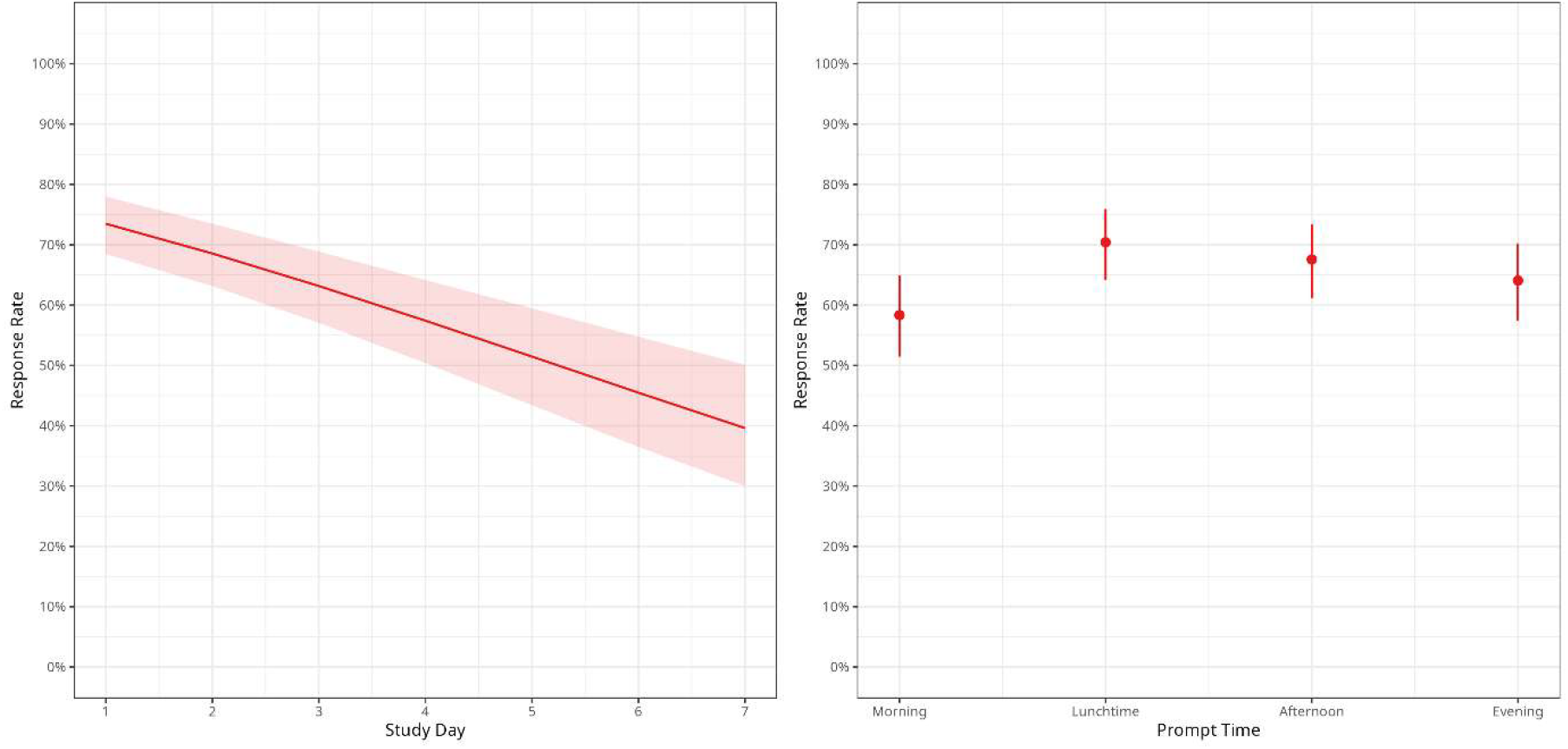
participants’ response rates against study day (left) and time of day (right). N=82.

The results of analyses estimating differences due to sex and age are shown in Figure 4. Girls responded more often to the μEMA prompts compared with boys (OR: 1.71 [95% CI: 1.03, 2.84]). However, the daily patterns were similar for both sexes (interaction term OR 1.07 [95% CI: 0.93, 1.23]). Response rate did not differ between age groups (OR: 0.73 [95% CI: 0.42, 1.27]) and daily response patterns were similar for the two age groups (study day by age interaction OR: 1.11 [95% CI: 0.95, 1.29]).

**Figure 4.**
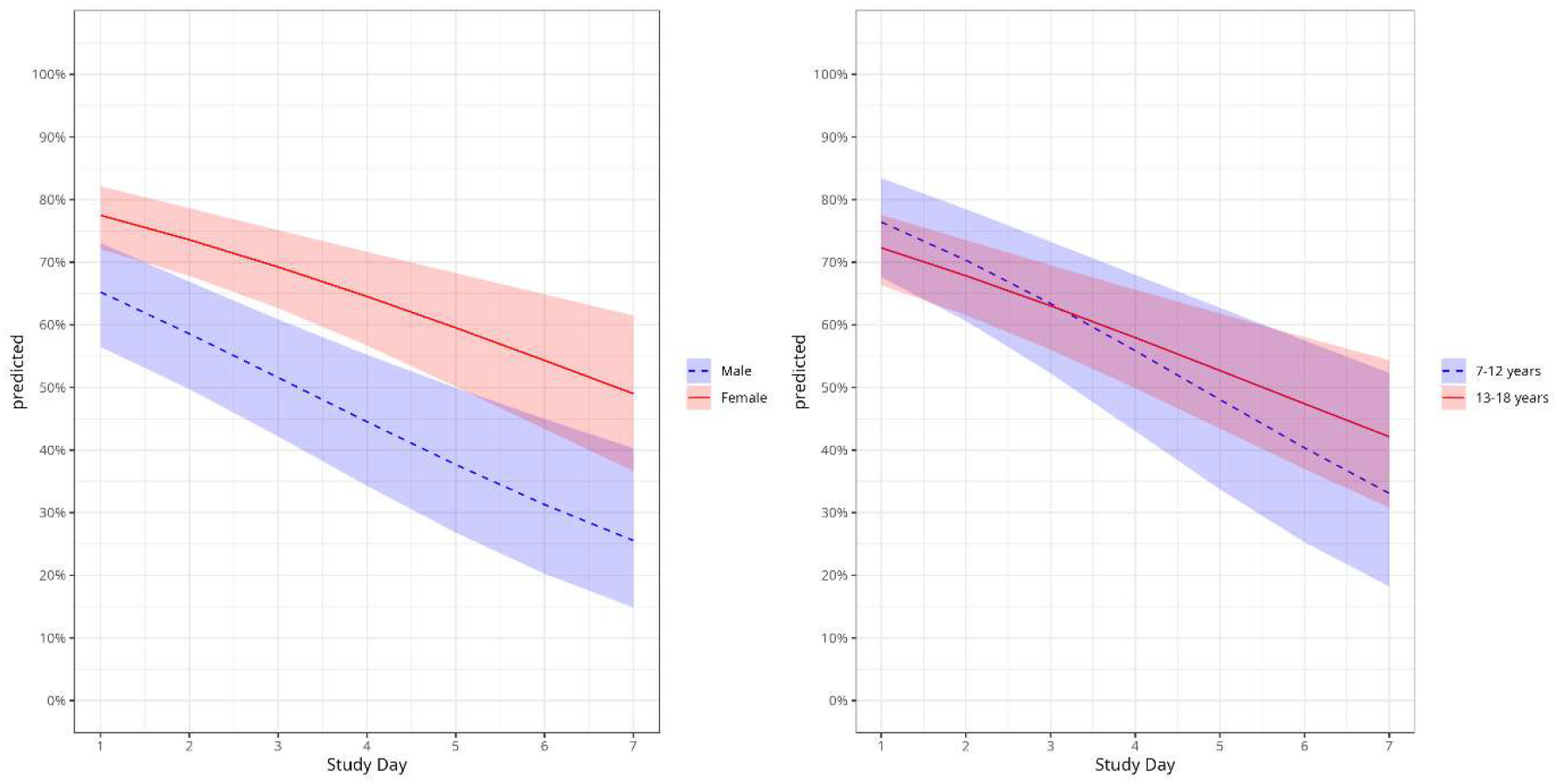
Association of μEMA prompt response with study day stratified by sex (left) and age (right).

### Evaluating participants survey responses on acceptability

A summary of responses to questions about smartwatch acceptability and wear time is provided in A total of 54 (65%) participants rated their experience using the smartwatch positively (a rating of 4 or 5 out of 5), 20 (24%) gave a neutral rating (3 out of 5) and 8 (10%) rated negatively (1 or 2 out of 5). In addition, 27 (33%) participants said they happy to participate in future studies using the smartwatch, while 39 (47%) said maybe, and 12 (14%) said no. The majority of participants who responded (n=61 [73.5%]) reported wearing the smartwatch for the entire duration of the study (8 days).

For those who reported that they did not wear the smartwatch for the entire duration of the study (22 participants), the most common reason was that they did not find it comfortable to wear (9/22, 41%). Other reasons included forgetting, that they did not see the benefit when they could not see the data, they were forbidden to wear it by school and it ran out of battery (all 3 or fewer responses).

## Discussion

### Principal Findings

In this feasibility study of a smartwatch based μEMA method to collect data on eating habits over 7 days in Malaysian children we found that most participants (67%) remained responsive to prompts up to the last day of the study. Participants were least likely to respond to prompts between 09:00-11:00, and most likely between 12:00-14:00. The intraclass correlation coefficient was 20.7%, suggesting that while some variation in response pattern is attributable to between-participant differences, the majority of the variation (79.3%) was due to within-participant differences. The response rate dropped off day-on-day and was higher for female than male participants; no association was found between participant age group and response rate.

Our average response rate of 69% was lower than the average of 78% found in a meta-analysis of EMA in children and adolescents, including studies that prompted between 2-9 times daily [18]. That study found that prompting participants more often had a large negative effect on completion rate, which is further supported by Kraft et al. [21] which found a negative correlation of -0.12 between increased number of prompts and response rate (p=0.009). Participants in our study were prompted 12 times a day, plus an additional catch-up prompt in the morning, which is likely to have had a negative effect on response rate, especially in the case of repeated “No food/drink” entries. It has been reported [18] in non-clinical studies that “a higher average compliance rate was observed in studies that prompted participants 2-3 times daily (91.7%) compared with those that prompted participants more frequently (4-5 times: 77.4%; 6+ times: 75.0%).” This suggests that compliance may be improved by prompting participants less frequently, for example by having three prompts daily at 11am, 3pm and 7pm although longer time intervals increase the reliance on memory potentially affecting the completeness of recorded consumption events. While on average, the (65%) participants rated the study protocol positively (either 4 or 5 out of 5), the response rate fell day-on-day. Participants were less likely to respond to prompts at the beginning or end of the day, compared with the middle of the day. Focus group or interview discussions were not feasible in this study due to COVID-19 restrictions, but should be explored in future studies to determine the reasons for missing event prompts and non-responses which may include forgetting or being involved in a competing activity when the prompt is sent [33].

Female participants had a higher response rate than male participants, consistent with previous findings [22] [34]. There was little evidence of difference in the relationship between response and study day for male versus female participants. An analysis of the SEACO-CH20 accelerometer dataset [35] found that a similar proportion of males and females had useable accelerometer data, suggesting that this was specific to the smartwatches rather than a difference with wrist-worn devices generally. Little evidence of a difference was found between response rates in the 7-12 and 13-18 age groups.

Although the subjective indicators suggested that most participants enjoyed wearing the smartwatch, only a minority of participants (33%) indicated that they would be willing to participate in a similar study again; 47% responded “Maybe”. Potential changes to the protocol that may improve compliance could include only wearing the smartwatch instead of both the smartwatch and accelerometer.

### Strengths and Limitations

This is the first study exploring the feasibility of using smartwatch-based EMA in a population of children and adolescents from a low-to-middle income country. This study was part of the SEACO study, using the SEACO-CH20 dataset, and lays the foundation for an improved understanding of the potential for wearable devices for measuring relationships between eating and cardiometabolic health. Data on 24-hour eating behaviours are important for informing policy that may reduce cardiometabolic risk among children and adolescents and prevent progression to cardiometabolic disease in adulthood; this study represents a move away from self-report after 24 hours and towards recording behaviour in real-time.

However, this study did have some limitations. The frequency of prompting may have affected compliance and should be discussed with participants to optimise the protocol for future studies in this population. It has been suggested [18] that compliance can be improved by incentivising participants with e.g. a monetary reward or raffle entries. Since this study concerns young people, one potential incentive method could be to gamify the EMA process using a level-up/promotion system in the app [36]. Previous studies have explored adding an end-of-day catchup prompt, which has been found to improve the reporting of dinner [33]. Replacing the morning catch-up prompt with one in the evening may improve response rate, especially given that we found that participants were more likely to respond to prompts in the evening than in the morning. Future studies may additionally consider using the catch-up entries to impute missed entries on the previous day, which could give more complete data. Another suggestion could be to incorporate a short period of training to improve response rates, where responses are monitored in real-time by researchers and participants are prompted directly by researchers if missing responses are common. Such an approach has been used to improve the accuracy of real-time food photography methods [37].

Unbalanced statistics limited our ability to assess differences across age, sex and ethnic group. The larger SEACO CH-20 accelerometer study[35] from which participants for this study were selected had more balanced statistics (49% female, 67% Malay, 44% <13 years old), which suggests that the cohort used for smartwatch data may not represent the overall SEACO CH-20 cohort. In particular, we only had 4 participants aged 7-9, so further studies are required to better understand the feasibility of dietary μEMA in the younger participants. Participants in this study began wearing the devices on different days of the week, and it is possible that day of the week could affect participation, for example whether it is a weekend or weekday – lower response rate at weekends has been previously documented by Battaglia [22]. We did not account for study start day due to the small sample size, and because schooling was disrupted throughout the study period due to the COVID-19 pandemic [38].

An issue with entry duplication meant that some entries may have been removed that were actual events not due to the software issue. This bug with the smartwatch software has since been fixed. Additionally, only the response time of the participant was recorded, and not the time that the prompt was sent. This means we had to infer which hour window the entries corresponded to – this could be programmed in the software.

Discomfort was the most common reason for non-wear cited by the participants, which may be unique to our study protocol that required participants to wear 2 wrist-worn devices on the same arm. Furthermore the smartwatch utilised in our study was not specifically designed to fit smaller children. Efforts to make smartwatches less intrusive, e.g. by making them smaller, may further improve response rate and study uptake.

Ramadan, a culturally important event in Malaysian society which includes fasting in some population groups, took place during the course of the data collection period. This is likely to have affected the eating behaviours of participants who took part during this time. To ensure that the number of EMA entries was not influenced by Ramadan, we excluded participants from our analyses who wore the smartwatch during the Ramadan period, thereby further limiting our sample size.

## Conclusions

This study extends previous eating behaviour studies by exploring the use of μEMA in a population of children and adolescents in Malaysia, and is the first such study to do so. Willingness to take part in the μEMA study was high but poor response rates suggest that the high number of prompts per day may be too burdensome. Further work is needed to explore different μEMA variations including using fewer prompts, and identify devices that may be more comfortable for child and adolescent participants. The growing use of smartwatches amongst children particularly in South-East Asia may offer more opportunities for further study [39].

## Abbreviations

μEMA: Micro-interaction Ecological Momentary Assessment
EMA: ecological momentary assessment
mEMA: mobile ecological momentary assessment
SEACO: South East Asia Community Observatory
SEACO-CH20: SEACO Child Health 2020

## Acknowledgements

For the purpose of open access, the author(s) has applied a Creative Commons Attribution (CC BY) licence to any Author Accepted Manuscript version arising from this submission. The Medical Research Council (MR/T018984/1) and the Ministry of Higher Education/UK-MY Joint Partnership on Non-Communicable Diseases (2019/MR/T018984/), both provided funding in support of this research. The SEACO health and demographic surveillance system is supported by Monash University. The study’s funders played no part in the study’s planning, gathering, analysing, or interpreting data, or in the report’s preparation. Sophia M. Brady is funded by the National Institute for Health and Care Research (NIHR) Applied Research Collaboration (ARC) North East and North Cumbria (NENC) (NIHR200173). The NIHR Bristol Biomedical Research Centre funds Miranda E.G. Armstrong (NIHR203315). The views expressed are those of the authors and not necessarily those of the NIHR or the Department of Health and Social Care. The authors also would like to express their appreciation to the SEACO Field Teams and survey participants. The South East Asia Community Observatory (SEACO, https://www.monash.edu.my/seaco) funded the research detailed in this paper. The authors’ opinions, however, are their own, and there is no real or implied sponsorship from SEACO.

## Data Availability

Data cannot be shared publicly for confidentiality and ethical reasons. De-identified data are available and can be freely requested from the South East Asia Community Observatory, Monash University Malaysia Institutional Data Access at ““ for researchers who meet the criteria for access to confidential data. For more information, please refer to https://www.monash.edu.my/seaco/research-and-training/how-to-collaborate-with-seaco.

