## Supplemental Material for "The Feasibility of Smartwatch Micro Ecological Momentary Assessment for Tracking Eating Patterns of Malaysian Children and Adolescents in the SEACO Child Health Update 2020: a Cross-Sectional Study"

Supplementary material

Methods

Data Collection

A flow chart of the original data collection plan can be seen below.

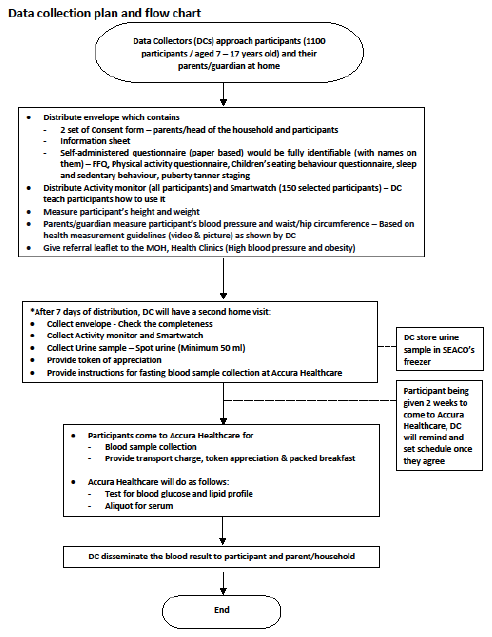

*Supplementary Figure 1 - the data collection plan.*

Smartwatch Prompts

The prompts sent to the smartwatches can be seen below. The prompts were sent in Malay; contact the corresponding author to request access to the Malay originals.

The regular prompts were worded as follows:

***English***

**Have you had any food or drink in the last hour?**

Yes (food or drink consumed in last hour)

No (no food or drink consumed in last hour)

**What did you have?**

Meal

Snack

Drink

*Back*

**What size was it?**

Small

Mid-size

Large

*Back*

**What did you use to eat?**

Hands

Fork/spoon

Chopsticks

*Back*

**Where were you?**

Home

School

Elsewhere

*Back*

**Any more food or drink to record?**

No

Yes

The wording of the catch-up prompt was:

***English***

**Any food or drink you forgot yesterday?**

No (no food or drink unrecorded from yesterday)

Yes (food or drink to be added from yesterday)

Results

Catch-up Entries

Survey

This section contains the text of the questions asked to participants (Supplementary Figure 2).

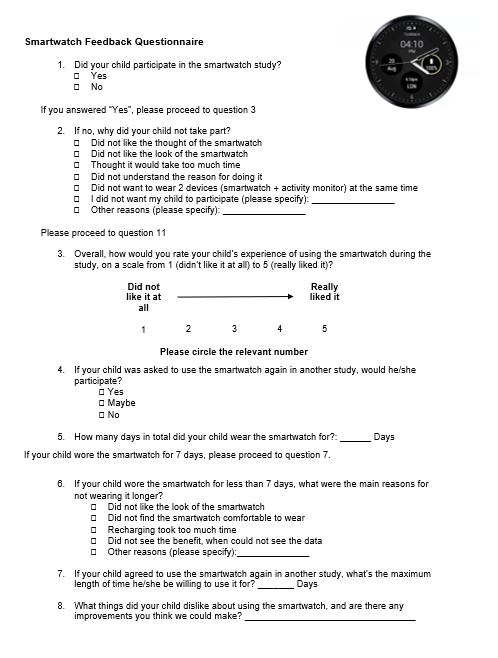

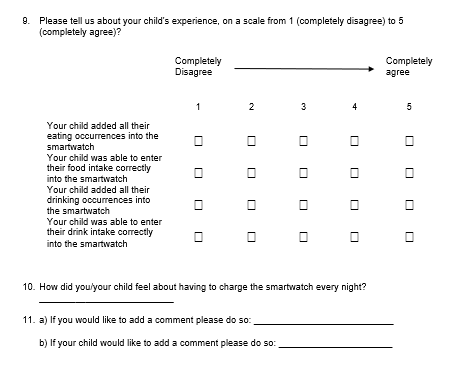

*Supplementary Figure 2 - the smartwatch survey. This version was given to the parents of the younger children; older children filled out their own surveys.*

*If you wore the smartwatch for less than 7 days, what were the main reasons for not wearing it longer?*

Six adolescents responded “Other” to this question and gave additional explanations. Translated into English, these are:

*Supplementary Table 1* - reasons that participants did not wear the watch for the duration of the study.

English

Forget

Forgotten

Forgot to use after swimming activity

The school does not allow the use of gadgets

Out of battery

Pain and itching

*If you agreed to use the smartwatch again in another study, what is the maximum length of time that you would be willing to use it for?*

A bar chart of participants’ responses can be seen in Supplementary Figure 3. The plurality of participants indicated that they would wear it for 7 or 8 days at most. The majority of participants indicated that they would wear it for at least as long as they had in this study. 8 participants refused to answer.

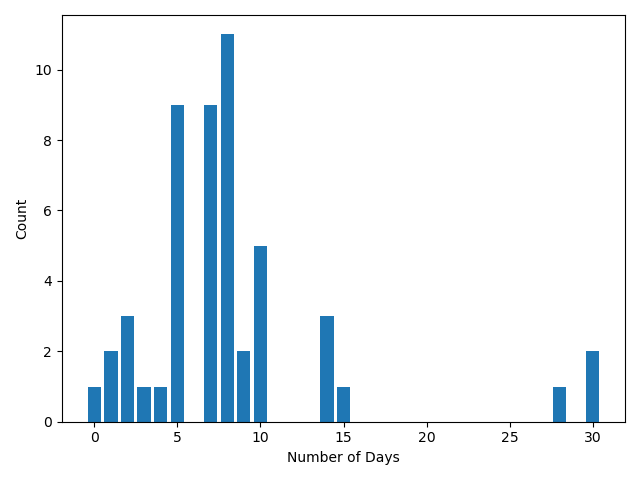

*Supplementary Figure 3 - the number of days that participants reported they would wear the smartwatch for in another study (N=59).*

*What things did you dislike about the smartwatch, and what improvements do you think we could make?*

Most participants provided no data in response to this question. Of those who did, translations of their responses are summarised in Supplementary Table 2.

*Supplementary Table 2* - what did you dislike about the smartwatch?

| Response (English) | Count |
| --- | --- |
| Tiada/Tidak | 7 |
| Semasa menulis/Tulis | 2 |
| 6 JAM SEHARI | 1 |
| Don’t use exercise time | 1 |
| Ask the same question | 1 |
| Placing the pattern | 1 |
| Heavy and easy to crack | 1 |
| There are improvements in the clock | 1 |

*Please tell us about your experience, on a scale of 1 to 5?*

The subjective experience for some aspects of the study are summarised in Supplementary Figure 4. The plurality of participants did not respond to these questions; of those who did, the majority felt that they entered all their eating occurrences and that they entered them correctly (either 4 or 5 on the scale). A large proportion of participants did not respond to this question, potentially indicating survey fatigue.

| Question | Response | N | % |
| --- | --- | --- | --- |
| 1. You added all your eating occurrences into the smartwatch | 1 (Completely disagree)  2 | 4 | 7 |
|  | 3 | 17 | 28 |
|  | 4 | 18 | 30 |
|  | 5 (Completely agree) | 22 | 36 |
| 2. You were able to enter your food intake correctly into the smartwatch | 1 (Completely disagree)  2 | 5 | 9 |
|  | 3 | 14 | 24 |
|  | 4 | 14 | 24 |
|  | 5 (Completely agree) | 25 | 43 |
| 3. You added all your drinking occurrences into the smartwatch | 1 (Completely disagree)  2 | 4 | 7 |
|  | 3 | 11 | 19 |
|  | 4 | 21 | 36 |
|  | 5 (Completely agree) | 22 | 38 |
| 4. You were able to enter your drink intake correctly into the smartwatch | 1 (Completely disagree)  2 | 4 | 7 |
|  | 3 | 13 | 22 |
|  | 4 | 17 | 29 |
|  | 5 (Completely agree) | 24 | 41 |

*Supplementary Figure 4 - responses to questions on their experience using the smartwatches (N=63).*

*How did you feel about having to charge the smartwatch every night?*

This was a free-text response, translations of the full responses can be seen in Supplementary Table 3.

*Supplementary Table 3* - participants' attitudes to charging the smartwatches (N=82).

| English | Count |
| --- | --- |
| Refused to answer | 14 |
| very comfortable | 1 |
| very good | 1 |
| very easy | 1 |
| best | 1 |
| Good | 3 |
| easy | 2 |
| easy to charge | 1 |
| Fun | 3 |
| Happy | 3 |
| happy | 2 |
| happy because i can use it tomorrow | 1 |
| easy | 2 |
| comfortable | 1 |
| comfortable | 1 |
| good | 1 |
| good and easy | 1 |
| quite easy | 1 |
| ok | 5 |
| ok | 1 |
| it's ok | 1 |
| ok | 1 |
| it's ok | 1 |
| ok | 1 |
| normal | 1 |
| it's normal | 1 |
| no feelings | 1 |
| none | 1 |
| uncomfortable | 1 |
| a little difficult | 1 |
| Easy | 1 |

*Any other comments?*

Translations of these can be seen in Supplementary Table 4. Most participants left no comment.

*Supplementary Table 4 - additional comments by the participants.*

| English | Count |
| --- | --- |
| 5 | 1 |
| 6 | 1 |
| Comfortable | 1 |
| It's fun | 1 |
| no | 1 |
| uncomfortable to wear because of wearing it for a long time | 1 |
| I feel really worried when wearing the monitor activity clock while sleeping for fear of damaging the clock due to knocking on the bedpost | 1 |

Smartwatch

Statistics on the median and IQR number of responses per day for all participants, including those who took part during Ramadan can be seen in Supplementary Table 5. Statistics for all participants including catch-up entries can be seen in Supplementary Table 6.

*Supplementary Table 5* - a summary of smartwatch responses by day, for all participants including those who took part during Ramadan.

| Median (IQR) | Meal | Drink | Snack |
| --- | --- | --- | --- |
| Day 1 | 2.0 (3.0) | 2.0 (4.0) | 1.0 (2.0) |
| 2 | 2.0 (2.0) | 2.0 (4.0) | 1.0 (2.0) |
| 3 | 2.0 (2.0) | 2.0 (3.0) | 0.0 (1.0) |
| 4 | 2.0 (2.0) | 1.0 (3.0) | 0.0 (1.0) |
| 5 | 1.0 (2.0) | 1.0 (2.0) | 0.0 (1.0) |
| 6 | 1.0 (2.0) | 1.0 (3.0) | 0.0 (1.0) |
| 7 | 0.5 (2.0) | 0.0 (2.0) | 0.0 (1.0) |

*Supplementary Table 6* - a summary of smartwatch responses by day for all participants, including catch-up events.

| Median (IQR) | Meal | Drink | Snack |
| --- | --- | --- | --- |
| Day 1 | 2.0 (3.0) | 2.0 (4.0) | 1.0 (2.0) |
| 2 | 2.0 (2.0) | 2.0 (4.0) | 1.0 (2.0) |
| 3 | 2.0 (2.0) | 2.0 (3.0) | 0.0 (1.0) |
| 4 | 2.0 (2.0) | 1.0 (3.0) | 0.0 (1.0) |
| 5 | 1.0 (2.0) | 1.0 (3.0) | 0.0 (1.0) |
| 6 | 1.0 (2.0) | 1.0 (3.0) | 0.0 (1.0) |
| 7 | 0.5 (2.0) | 0.0 (2.0) | 0.0 (1.0) |

The median of the mean response rate per day is shown in Supplementary Table 7.

Supplementary Table 7 - the median and interquartile range of response rates per day.

| Day | Median /% | IQR |
| --- | --- | --- |
| 1 | 83 | 67, 92 |
| 2 | 75 | 58, 92 |
| 3 | 75 | 50, 83 |
| 4 | 67 | 48, 83 |
| 5 | 75 | 42, 84 |
| 6 | 67 | 42, 83 |
| 7 | 58 | 33, 75 |
